# Progastrin, Annexin A2 and Tumor-Associated Macrophages in Gastric Adenocarcinomas

**DOI:** 10.1101/2025.09.10.25334872

**Authors:** Konstantinos Christofidis, Rodanthi Fioretzaki, Stylianos Mavropoulos Papoudas, Nikolaos Charalampakis, Nikolaos Kavantzas, Dimitrios Schizas, Stratigoula Sakellariou

## Abstract

**Introduction:** This study investigates the immunohistochemical expression of hPG, ANXA2, and the polarization of tumor-associated macrophages (TAMs) in gastric adenocarcinomas. Methods: A retrospective analysis was conducted on FFPE tissue samples from 60 patients with gastric adenocarcinoma (primary tumors, lymph node metastases and normal-looking gastric mucosa), and on gastric biopsies from 23 healthy controls. The expression of hPG and ANXA2 was quantified using the H-score, and the CD163/HLA-DR ratio was used to infer macrophage polarization (M2/M1). Results: ANXA2 expression was significantly elevated in primary tumors and lymph node metastases compared to normal and healthy controls and increased with tumor grade. High ANXA2 expression was associated with poorer overall and disease-free survival. In contrast, hPG expression, although positively correlating to ANXA2 expression, showed no prognostic value. The M2/M1 ratio increased with tumor progression, showed a negative correlation with ANXA2 expression and failed to correlate significantly with survival. Conclusions: This study is the first to demonstrate the adverse prognostic impact of ANXA2 overexpression in gastric adenocarcinoma tissues from Caucasian patients, hinting to its potential utility as a prognostic biomarker and therapeutic target. Further large-scale studies could aid validate these findings and explore the therapeutic potential of targeting ANXA2 and modulating the TAM polarization.

## Introduction

### 1.1 Gastric Adenocarcinoma

Gastric cancer is the fifth most common cancer globally, with 968,350 new cases and 659,853 deaths annually. Its incidence rises with age, with the average age of diagnosis being 70 years. Gastric cancer is anatomically classified into two subtypes: cardia and non-cardia. Chronic *Helicobacter pylori* infection is the main cause of 90% of non-cardia gastric cancers, while additional risk factors include diet, alcohol, smoking, and Epstein-Barr virus infection. Importantly, *Helicobacter pylori* eradication treatments, improved nutrition and hygiene have led to an incidence decline of this particular type of gastric cancer. Cardia cancers are less associated with *Helicobacter pylori* (20%) and more often linked to obesity and gastroesophageal reflux. Their incidence tends to increase in the younger population. [1–3] A family history, particularly of hereditary diffuse gastric cancer caused by mutations in the cadherin 1 gene, accounts for less than 10% of cases. [4,5]

Gastric adenocarcinomas are histologically classified according to the WHO 2019 guidelines into tubular, poorly cohesive (including signet-ring cell carcinoma), and mixed adenocarcinomas. Rare subtypes include the papillary, mucinous, hepatoid, micropapillary and fundic-gland type adenocarcinomas, the carcinoma with lymphoid stroma, as well as the mucoepidermoid, Paneth cell and parietal cell carcinomas. [6] Despite progress in medicine, early detection methods, sensitive biomarkers and effective therapies are still lacking. [7,8] As a result, active research is ongoing among many molecules implicated in gastric cancer oncogenesis.

### 1.2 Progastrin

Progastrin (hPG), an 80-amino-acid precursor of amidated gastrin, is synthesized in gastric antral G cells [9,10]. Normally, the non-amidated gastrins make up <10% of the total secreted peptide forms. Elevated levels are seen in pathological states, including cancers [11,12], due to *GAST* gene overexpression on chromosome 17q21 [11,13,14] and deficient processing enzymes in tumors [15–17]. hPG has been shown to promote cancer cell proliferation [18], resistance to apoptosis [19], disruption of cell junctions [20], and supports cancer stem cell properties [18,21] and angiogenesis. [22] Moreover, hPG suppresses M2 macrophage polarization and Wnt ligand secretion. [23] hPG acts via multiple pathways, including Wnt/β-catenin, KRAS, MEK-ERK, PI3K/Akt, NF-κB, and SMAD4 [24], and is found in tumor cells and stroma. [25] Increased hPG levels found in samples of patients with 11 different types of cancer, hints to its potential value as a biomarker. [11] The majority of the studies have examined serum hPG levels [26–36], with only a few studies researching its expression in tumor tissues. [37–40] When it comes to the role of hPG in gastric adenocarcinomas, limited research has been carried out up to this date, mainly using cell lines and mouse models. [41–43] The receptors for hPG are still unidentified, though Annexin A2 is a proposed candidate. [44–45]

### 1.3 Annexin A2

Annexin A2 (ANXA2) is a 36-kDa phospholipid-binding protein encoded by the *ANXA2* gene on chromosome 15q22.2. [46–48] It regulates multiple cellular functions and is implicated in tumorigenesis through pathways involving c-Myc, STAT3, SNAIL, TWIST, ARP3, MIEN1, LIMK and CFL1. [49] ANXA2 also seems to be critical for hPG’s oncogenic action, at least in colon cancer cells. [45,50] In gastric cancer, ANXA2 is overexpressed and is localized mainly at the tumor cell membranes. Among other effects, its overexpression is linked to destabilization of epithelial junctions and increased matrix metalloproteinases’ secretion. Tumor cells with high ANXA2 expression have an increased invasive and metastatic potential [51–55]. ANXA2 is also linked to *c-erbB-2* overexpression and poor patients’ outcomes. [56] Furthermore, the ANXA2 axis has emerged as a potential therapeutic target as its silencing restrains tumor cell proliferation and survival and reverses chemoresistance. [57–60]

### 1.4 Tumor-Associated Macrophages

Tumor-Associated Macrophages (TAMs) are a critical component of the tumor microenvironment and play a pivotal role in tumor progression. They are immune cells that, following various stimuli, polarize, creating a multitude of activated macrophages whose properties and functions vary. Activated macrophages are classified into the M1 phenotype that induces inflammation and shows microbicidal and tumor suppressive activity, and the M2 phenotype, with immunoregulatory activity that contributes to tissue healing, but also to cancer development, with many more phenotypes existing between these two ends of the spectrum. M1 markers include HLA-DR, CD86, iNOS, and pSTAT1, while M2 markers include CD163, CD204, and CD206. [61] M1 polarization is induced by TLR ligands, TNF-α, IFNγ, and CSF2, while IL4, IL10, IL13, TGF-β, and PGE2 promote M2 differentiation. [62] A high M2/M1 TAM ratio is associated with worse prognosis in several cancers [63], including gastric cancer [64–68] and has been linked to some aspects of the gastric adenocarcinoma oncogenesis. [69–71] As previously mentioned, hPG has been reported to suppress the differentiation of TAMs towards the M2 phenotype and reduce the expression of Wnt ligands in them. [23] It is also interesting that ANXA2 is expressed on the surface of macrophages, serving as a recognition element and mediating their activation. [72]

This study investigates the immunohistochemical expression of hPG, ANXA2 and the phenotype of TAMs in gastric adenocarcinoma patients, aiming to reveal a possible interplay among these molecules, as well as any existing associations with prognostic factors in order to shed light in gastric cancer tumorigenesis and propose new prognostic biomarkers.

## 2. Materials and Methods

### 2.1. Patients and tissue samples

This is a retrospective study carried out on gastrectomy specimens from 60 patients who underwent surgery for gastric adenocarcinomas at the First Department of Surgery, National and Kapodistrian University of Athens (NKUoA), “Laiko” University Hospital between 2014 and 2020. For these patients, a complete dataset was available, including a prospective recording of their demographic data, type of surgery, TNM histological classification, pre- and postoperative chemotherapy, as well as their postoperative follow-up, including their potential recurrence, their disease-free and overall survival. A summary of this data is shown in Table 1.

**Table 1.** Clinicopathological characteristics of patients.

| Parameter | Median | Range or % |
| --- | --- | --- |
| **Age (years)** | 67 | 34–86 |
| **Gender** |  |  |
| Male | 35 | 58.3% |
| Female | 25 | 41.7% |
| **Location** |  |  |
| Non-cardia | 42 | 60% |
| Cardia | 18 | 40% |
| **Surgical Procedure** |  |  |
| Subtotal Gastrectomy | 33 | 55% |
| Total Gastrectomy | 27 | 45% |
| **Histological Subtype** |  |  |
| Tubular | 27 | 45% |
| Poorly Cohesive | 24 | 40% |
| Mixed | 7 | 11.7% |
| Mucinous | 2 | 3.3% |
| **Chemotherapy** |  |  |
| No | 51 | 85% |
| Yes | 9 | 15% |
| **T category** |  |  |
| T1 | 9 | 15% |
| T2 | 6 | 10% |
| T3 | 22 | 36.7% |
| T4 | 23 | 38.3% |
| **N category** |  |  |
| N0 | 15 | 25% |
| N1 | 10 | 16.7% |
| N2 | 13 | 21.7% |
| N3 | 22 | 36.6% |
| **Grade** |  |  |
| 1 | 0 | 0% |
| 2 | 17 | 28.3% |
| 3 | 43 | 71.7% |
| **Stage** |  |  |
| IA | 6 | 10% |
| IB | 3 | 5% |
| IIA | 7 | 11.7% |
| IIB | 4 | 6.7% |
| IIIA | 8 | 13.3% |
| IIIB | 16 | 26.6% |
| IIIC | 6 | 10% |
| IV | 10 | 16.7% |
| **Event** |  |  |
| Death of Disease | 37 | 61.7% |
| Remission | 3 | 5% |
| Free of Disease | 20 | 33.3% |
| **Survival (days)** |  |  |
| Overall | 1374 | 149-3413 |
| Disease-Free | 984.5 | 100-3413 |

Formalin-fixed, paraffin-embedded tissue samples from the above-mentioned patients were retrieved from the archives of the Pathology Department of the same hospital. Sections from the primary site of gastric adenocarcinomas and, when applicable, their lymph node metastases were studied. Two control groups were included: non-tumoral/normal-looking gastric mucosa adjacent to the carcinomas derived from the same patients’ surgical specimens, as well as lesional-free gastric biopsies from 23 age and sex matched healthy subjects without any history of gastric cancer. All cases were anonymized, and each sample was assigned an alphanumeric code, in order to ensure the protection of the identity of the individuals. Permission for scientific use of patient data was obtained by the Research Ethics and Deontology Committee, NKUoA (492/18-07-2022). Individual consent was waived due to the nature of the study.

### 2.2. Immunohistochemical staining and evaluation

Immunohistochemistry (IHC) was performed on 3-4 µm-thick tissue sections according to standard procedure, which were then stained with commercially available rabbit monoclonal IgG antibodies against HLA-DR (clone EP96 at 1:200 dilution, Bio SB, Santa Barbara, CA, U.S.A.), CD163 (clone D6U1J at 1:250-1:1000 dilution) and ANXA2 (clone D11G2 at 1:200-1:800 dilution) (Cell Signaling Technology, Inc., Danvers, MA, U.S.A.). For hPG, there were no commercially available antibodies. A rabbit polyclonal antibody (1137 at 1:1000 dilution) was kindly provided by Professor Arthur Shulkes and his research team at the University of Melbourne, Victoria, Australia.

IHC evaluation to assess the expression of hPG, ANXA2, CD163 and HLA-DR at the protein level was performed by two surgical pathologists (K.C. and S.S.) who were blinded to clinicopathological information. Evaluation was performed by both pathologists simultaneously using a double-headed-microscope. At least 10 HPFs were scanned for each IHC stain for every case. The scoring system for hPG (cytoplasmic staining) (Figure 1) and ANXA2 (membranous staining) (Figure 2) was set based on the percentage of the stained tumor cells (0-100% of positive tumor cells) and the intensity of the immunostain (0: no staining; 1: weak; 2: moderate; 3: strong). We calculated the H-score for these two immunostains, by the following formula:

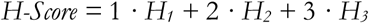

**Figure 1.**
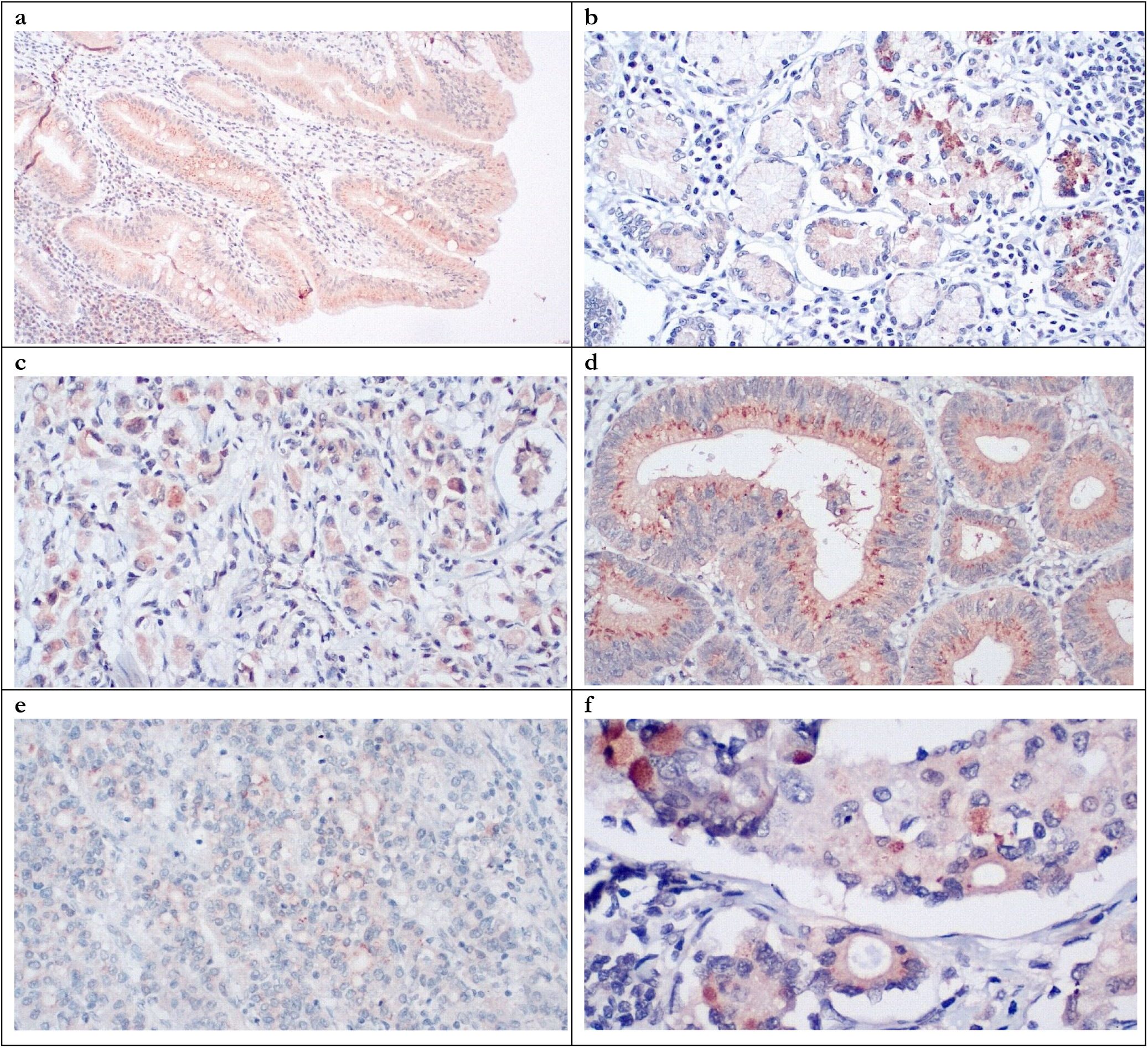
hPG expression in Normal Tissue (a, b - moderate to strong expression), Primary Tumor (c, d - weak to strong expression) and Lymph Node Metastasis (e, f - moderate to strong expression). Note the variety and heterogeneity of staining intensity even among similar looking cells of the same area.

**Figure 2.**
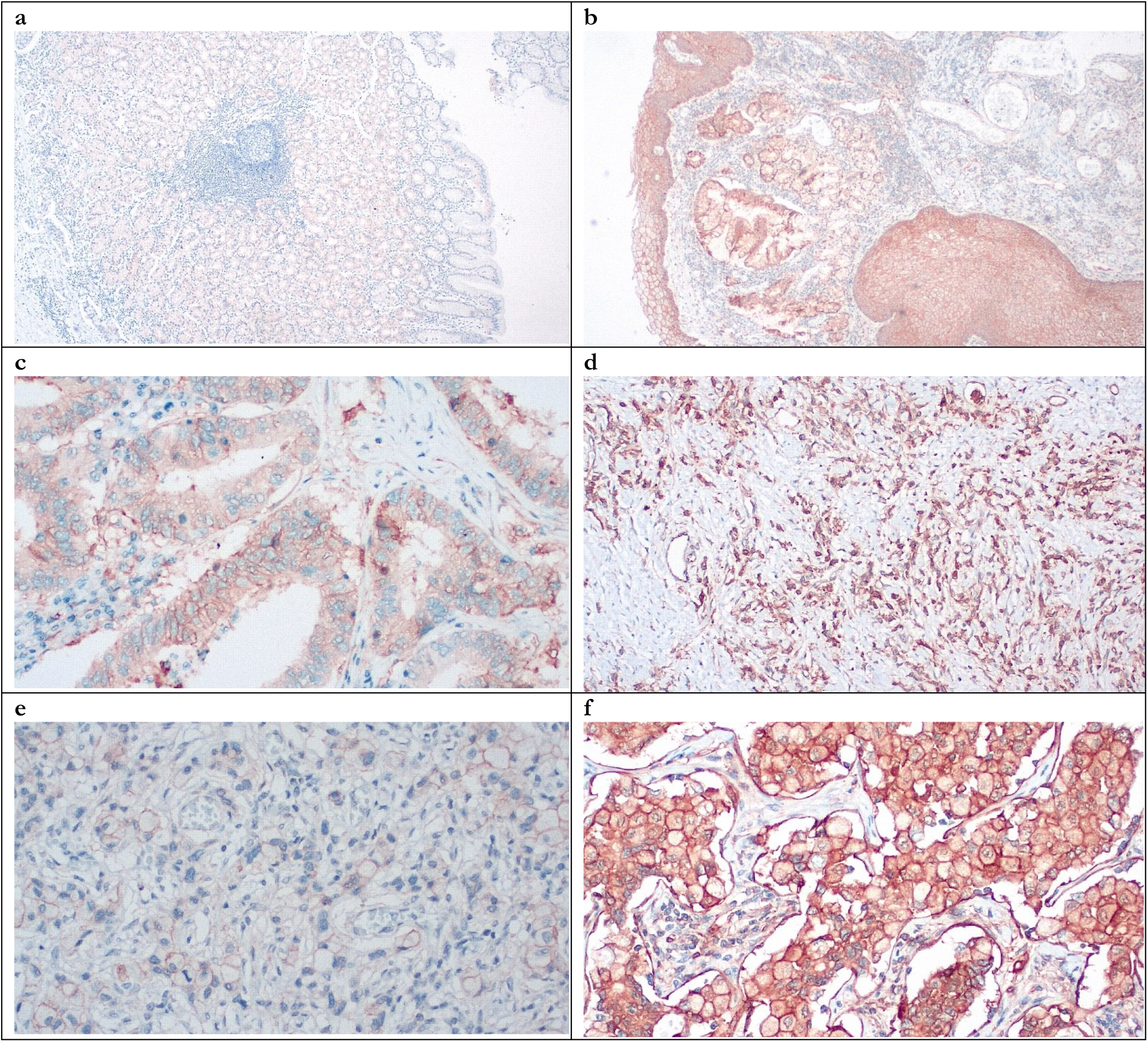
ANXA2 expression in Normal Tissue (a, b - weak to strong expression), Primary Tumor (c, d, moderate to strong expression) and Lymph Node Metastasis (e, f - weak to strong expression).

In this formula, H_1_, H_2_ and H_3_ represent the percentage of tumor cells showing weak, moderate and strong immunostaining, respectively. Since the maximum percentage of cells positive for any immunostain is 100%, H-score values ranged from 0-300.

Considering the distribution of values in the dataset, a threshold of 100 was applied to compare survival groups. H-score values < 100 signify low expression, whereas h-score values ≥ 100 signify high expression for these immunostains.

When evaluating CD163 and HLA-DR, the total number of positive macrophages was calculated for each immunostain per 10 HPFs (Figure 3). The ratio of CD163/HLA-DR was assessed, with values greater than 1 signifying a predominance of M2 compared to M1 macrophages.

**Figure 3.**
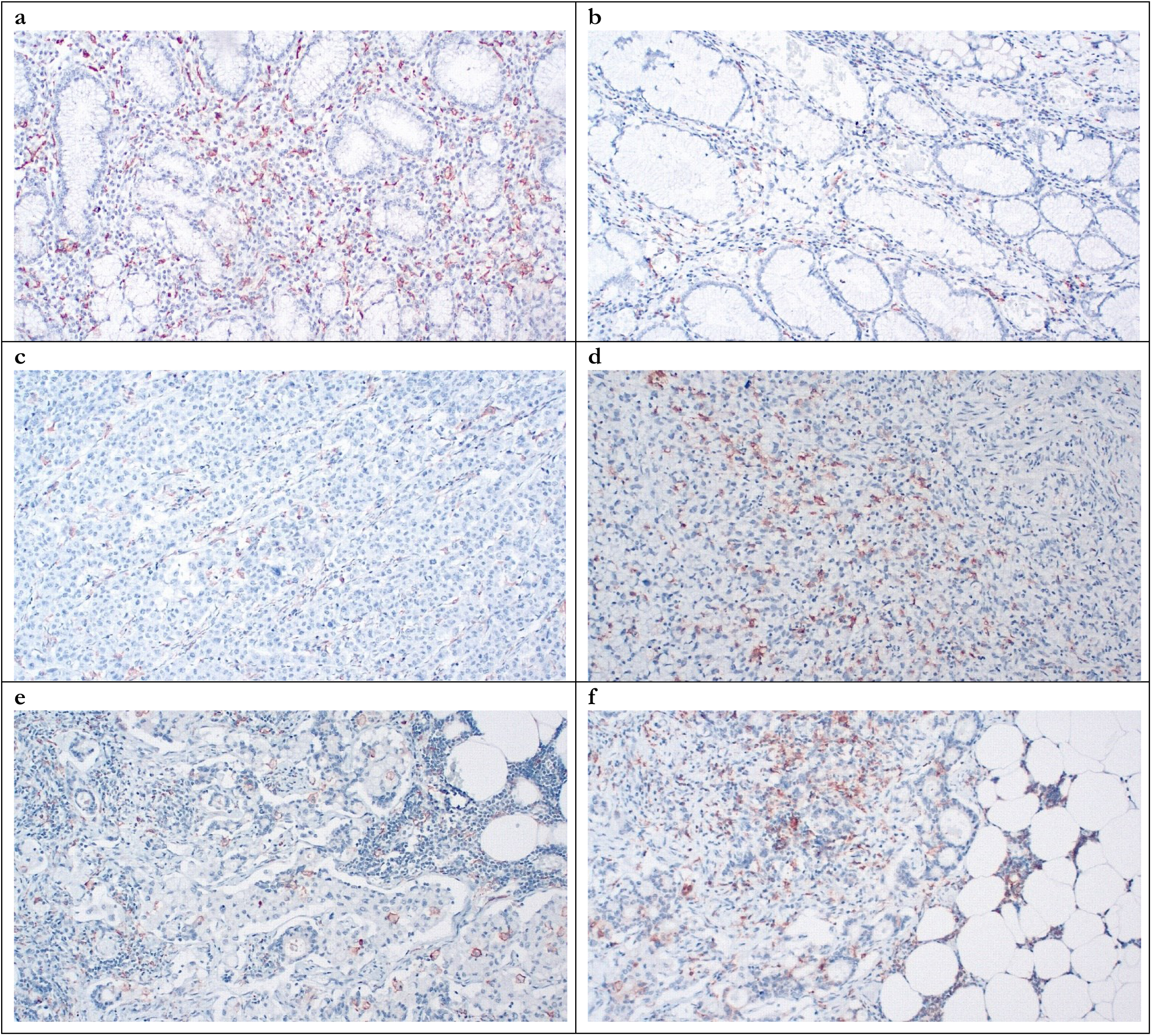
HLA-DR (a, c, e) and CD163 expression (b, d, f) in Normal Tissue (a, b), Primary Tumor (c, d) and Lymph Node Metastasis (e, f).

### 2.3. Statistical analysis

Statistical analysis was performed using R version 4.1.2 (2021-11-01). Data were expressed as frequencies, mean with SD or median with interquartile range (IQR), as appropriate. Quantitative variables were compared with Student’s t test or Mann–Whitney test for normally distributed and non-normally distributed variables, respectively. Qualitative variables were compared with the Chi-squared test or Fisher’s exact test, as appropriate. Relationships between parameters were assessed using Spearman’s correlation coefficient. All tests were two-sided and p values < 0.05 were considered significant.

To investigate IHC expression differences, we applied a non-parametric Wilxocon test for paired samples, when dealing with samples from the same patient, or for unpaired samples, when the samples came from different patients. Correlations between the expression of hPG, ANXA2 and the CD163/HLA-DR ratio in the tumor or the lymph node metastasis were investigated by Spearman’s rank correlation coefficient (ρ). A p-value was calculated to determine statistical significance. Relationships between the expression of hPG, ANXA2, as well as the CD163/HLA-DR ratio and the patients’ clinicopathological parameters (T, N, Stage, Grade and Histological Subtype) were assessed using a Kruskal-Wallis ANOVA test. The Dunn test was used to assess the Kruskal-Wallis ANOVA ability to differentiate between subgroups. As already mentioned, for the statistical analysis of the hPG and ANXA2 immunostaining expression, H-score values <100 were considered low expression, whereas values ≥ 100 high expression. For the CD163/HLA-DR ratio, values <1 show a greater influence of HLA-DR, since it is located in the denominator and values ≥1 point to the numerator, CD163, being more influential. Finally, we investigated the different projected survival outcomes for overall and disease-free survival regarding the expression of hPG and ANXA2, as well as the CD163/HLA-DR ratio in the tumor and the lymph node metastases. Survival curves were estimated using the Kaplan-Meier method, and differences between groups were compared using the log-rank test to obtain a p-value.

## 3. Results

Evaluation of immunohistochemical hPG, ANXA2, CD163 and HLA-DR expression was feasible in all tumoral [primary gastric tumors (T) and lymph node metastases (LN)] and non-tumoral tissues [normal-looking gastric tissues of the same patients (“normal”) and healthy controls (“healthy”)]. Concerning hPG, the staining pattern was cytoplasmic, indicative of the protein location. H-score ranged between 0 and 159 in T samples, 0 and 166 in LN samples, 8 and 157 in patients’ normal tissues, and 35-160 in healthy controls. For ANXA2, the staining pattern was membranous, supportive of its role as a receptor. H-score ranged between 0 and 242 in T samples, 0 and 285 in LN samples, 3 and 166 in patients’ normal tissues, and 5-107 in healthy controls. CD163/HLA-DR ratio ranged between 0,07442 and 6,95161 in T samples, 0,1502 and 4,2667 in LN samples, 0,1948 and 3,3095 in patients’ normal tissues, and 0,1159-1,0545 in healthy controls. The summary distributions of the hPG, ANXA2 and CD163/HLA-DR values in the various tissues examined are depicted in Suppl. Table 1. Both ANXA2 and the CD163/HLA-DR ratio seem to be rising when we move from “healthy” to “normal” to tumor tissue and to lymph node metastases, whereas hPG expression decreases (Figure 4).

**Figure 4.**
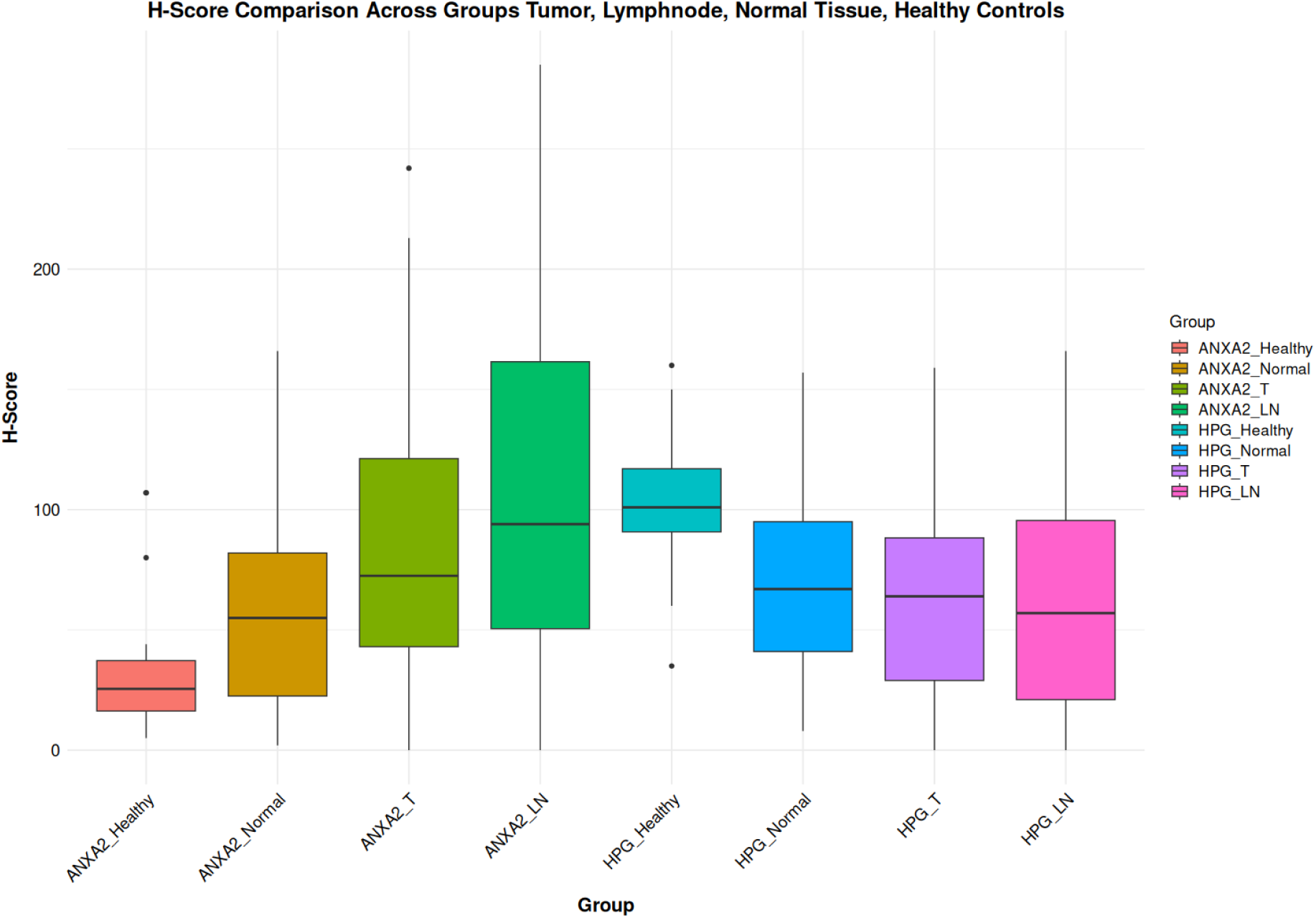
Comparison of the hPG and ANXA2 H-Score values in the gastric mucosa of healthy controls (Healthy), patients’ normal gastric mucosa (Normal), primary tumor (T) and lymph node metastases (LN).

### 3.1. hPG, ANXA2, CD163/HLA-DR differential expression

As hinted by the summary distributions, hPG, ANXA2, CD163/HLA-DR differs between tumoral (T and LN) and non-tumoral tissues (“normal” and “healthy”). We continued the analysis by researching the statistical importance of these expression differences. All the tests carried out concluded in statistically significant differential expressions, except the one between hPG (T) or hPG (LN) and the “normal” gastric tissues of the same gastrectomy specimen.

We also tested the markers’ differential expression between the two kinds of control tissues, “normal” and “healthy”. These tests also highlighted the statistically important differential expressions, except the one comparing the CD163/HLA-DR ratio between the “normal” and “healthy” controls. These results are shown in Table 2.

**Table 2.** Differential expression of hPG, ANXA2 and the CD163/HLA-DR ratio between the primary tumor (T) or the lymph node metastases (LN) and the patients’ normal gastric mucosa (Normal) or healthy controls (Healthy).

|  | p-value |
| --- | --- |
| hPG (T) vs hPG (Normal) | 0,3435 |
| hPG (T) vs hPG (Healthy) | **0,00000000005275** |
| hPG (Normal) vs hPG (Healthy) | **0,0005055** |
| hPG (LN) vs hPG (Normal) | 0,9751 |
| hPG (LN) vs hPG (Healthy) | **0,00000005451** |
| ANXA2 (T) vs ANXA2 (Normal) | **0,00007515** |
| ANXA2 (T) vs ANXA2 (Healthy) | **0,00000000003586** |
| ANXA2 (Normal) vs ANXA2 (Healthy) | **0,02435** |
| ANXA2 (LN) vs ANXA2 (Normal) | **0,001125** |
| ANXA2 (LN) vs ANXA2 (Healthy) | **0,00000002517** |
| CD163/HLA-DR (T) vs CD163/HLA-DR (Normal) | **0,000002619** |
| CD163/HLA-DR (T) vs CD163/HLA-DR (Healthy) | **0,00007961** |
| CD163/HLA-DR (Normal) vs CD163/HLA-DR (Healthy) | 0,1077 |
| CD163/HLA-DR (LN) vs CD163/HLA-DR (Normal) | **0,0002054** |
| CD163/HLA-DR (LN) vs CD163/HLA-DR (Healthy) | **0,0005706** |

### 3.2. hPG, ANXA2, CD163/HLA-DR expression correlation

As can be seen in Table 3, we also proved that the expression of hPG, ANXA2 and CD163/HLA-DR in the primary tumor is linked to each marker’s expression in the corresponding lymph node metastasis. More precisely, there exists a statistically significant positive correlation, meaning that the higher the expression in the primary tumor, the higher it will be in the lymph node metastasis as well. We also investigated the correlations between the different markers’ expression, both in the primary tumors as well as the lymph node metastases. A statistically significant positive correlation was proven to exist between ANXA2 and hPG expression in the primary tumors. Finally, a statistically significant negative correlation was demonstrated between the expression of ANXA2 and the CD163/HLA-DR ratio in the tumor tissues (Table 4). No other expression correlation came to light by the rest of the tests performed.

**Table 3.** Correlation of the expression of hPG, ANXA2 and the CD163/HLA-DR ratio in the primary tumor (T) versus the lymph node metastases (LN).

| Correlation | p-value | rho |
| --- | --- | --- |
| hPG (T) vs hPG (LN) | **0,00002913** | 0,5918522 |
| ANXA2 (T) vs ANXA2 (LN) | **0,00002263** | 0,5982855 |
| CD163/HLA-DR (T) vs CD163/HLA-DR (LN) | **0,00001069** | 0,6166044 |

**Table 4.** Correlation of the expression of hPG versus ANXA2 versus the CD163/HLA-DR ratio in the primary tumor (T) or the lymph node metastases (LN).

| Correlation | p-value | rho |
| --- | --- | --- |
| hPG (T) vs ANXA2 (T) | **0,0009792** | 0,4149362 |
| hPG (T) vs CD163/HLA-DR (T) | 0,3781515 | -0,1158293 |
| hPG (LN) vs ANXA2 (LN) | 0,8053 | 0,03872309 |
| hPG (LN) vs CD163/HLA-DR (LN) | 0,9599 | -0,007895732 |
| ANXA2 (T) vs CD163/HLA-DR (T) | **0,04332532** | -0,2617916 |
| ANXA2 (LN) vs CD163/HLA-DR (LN) | 0,6521 | 0,07075436 |

### 3.3. hPG, ANXA2, CD163/HLA-DR expression in relation to clinicopathological parameters

Possible relationships between the expression of hPG, ANXA2, as well as the CD163/HLA-DR ratio and the patients’ clinicopathological parameters (T and N categories of TNM, Stage, Grade and Histological Subtype) were researched. We found a relationship between the tumor’s grade and the expression of ANXA2 in the lymph node metastases, as well as the CD163/HLA-DR ratio, both in the tumor and the lymph node metastases (Suppl. Table 2). In the case of ANXA2 (LN), its expression seems to increase as the Grade progresses from 2 to 3 (p-value: 0,016). Reversely, the CD163/HLA-DR (T) and (LN) ratio appears to decrease with increasing tumor grade (p-values 0,04 and 0,02) (Suppl. Table 3). As mentioned in the statistical analysis section, the Dunn test was used to assess the Kruskal-Wallis ANOVA ability to differentiate between subgroups. The above-mentioned results were retained by the Dunn test, as our database contained only Grade 2 and 3 tumors.

A correlation was also shown between hPG (T) expression and the tumor stage, between hPG/ANXA2 (T) and the N parameter of the TNM, as well as between hPG/ANXA2 (LN) and the T parameter of the TNM. However, according to the Dunn test, in the first correlation, the test appears to only differentiate the expression of hPG (T) between Stages I and II, whereas the database comprised of tumors of all possible stages (I-IV). Therefore, the difference in hPG (T) expression was not significant when comparing Stages I and III, I and IV, II and III, II and IV, III and IV. Likewise, for the second correlation, the test only differentiates significantly between N values of 2 and 3 and between T subgroups of 2 and 3 for the third correlation, whereas more subgroups of these parameters exist. These results indicate a failure to properly differentiate these patients’ clinicopathological parameters’ values based on the expression of these antigens and thus these correlations were discarded (Suppl Table 2).

### 3.4. Marker expression and projected patients’ survival outcomes

Finally, we investigated the different projected survival outcomes for overall and disease-free survival regarding the expression of all markers in tumoral tissues. The expression of ANXA2, both in the tumor and the lymph node metastases, was the one to show statistically significant different survival curves, with a worse survival outcome being associated with higher ANXA2 expression. This negative relationship between ANXA2 expression and patients’ survival can be seen in Suppl. Table 4 and Figures 5-6, where overall survival and disease-free survival are severely impacted when ANXA2 expression is higher. Patients with high ANXA2 expression in the lymph node metastases showed the worst prognosis. The survival analysis results for ANXA2 have also been plotted in combined plots, comparing the different survival groups (Figures 5-6). As such, our population is divided into two distinct strata, patients with ANXA2 H-score values < 100, and patients with ANXA2 H-score values ≥ 100.

**Figure 5.**
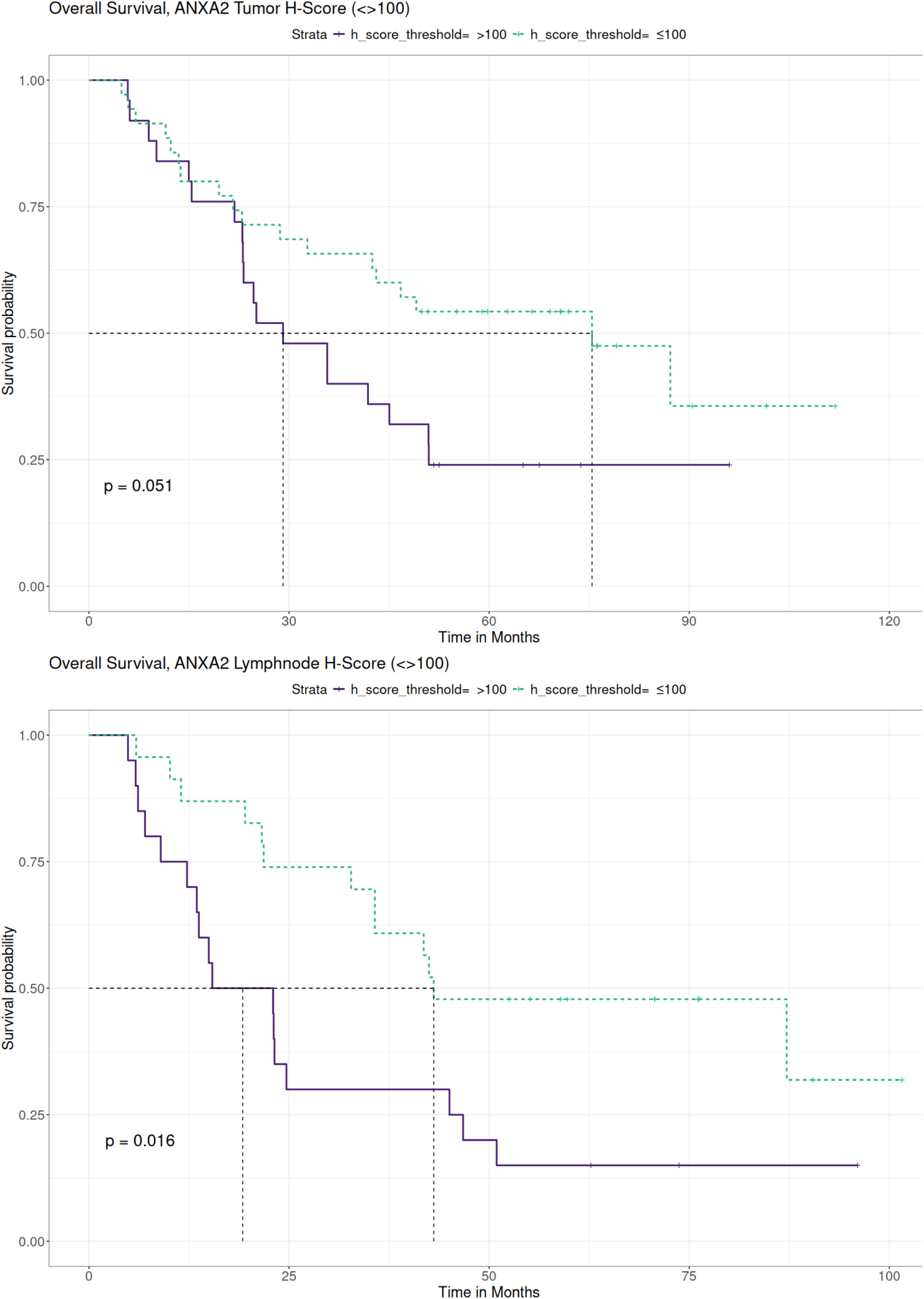
Comparison of Survival Curves depicting patients’ overall survival stratified by high and low ANXA2 expression in the tumor and the lymph node metastases. Survival was worse for patients with high ANXA2 expression.

**Figure 6.**
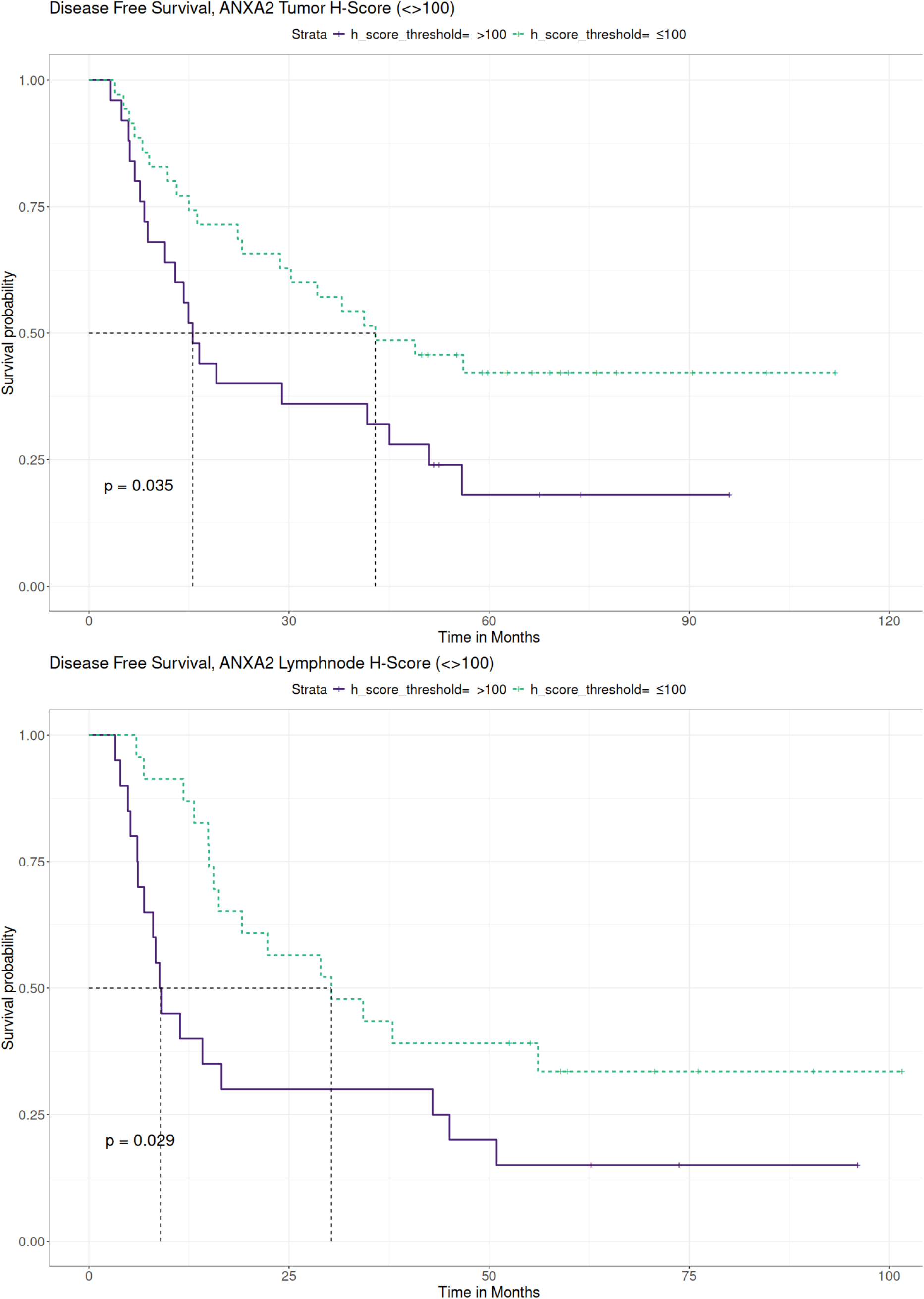
Comparison of Survival Curves depicting patients’ disease-free survival stratified by high and low ANXA2 expression in the tumor and the lymph node metastases. Survival was significantly worse for patients with high ANXA2 expression.

For hPG, a higher expression seemed to coincide with better patients’ survival outcomes. However, as shown in Suppl. Table 5, all the tests have p-values higher than 0,05 making them statistically non-significant. Consequently, patients’ overall and disease-free survival were not influenced by hPG expression. Likewise, the ratio of CD163/HLA-DR was assessed for differences in survival. The threshold of 1 was used, with values <1 showing a greater influence of HLA-DR, and values ≥1 pointing to CD163 being more influential. Although the ratio seemed surprisingly to be higher in patients with better survival outcomes, these results were not statistically significant (p-values >0,05).

## 4. Discussion

Gastric adenocarcinoma remains a major global health problem worldwide. Despite the advances in medicine, the prognosis for advanced-stage gastric cancer remains poor. In this retrospective study, we studied hPG, ANXA2, and the TAMs, aiming to determine their potential roles in gastric cancer progression and their prognostic significance. We therefore investigated their expression in gastric adenocarcinomas (T) and their lymph node metastases (LN), in non-tumoral gastric tissue adjacent to adenocarcinomas (“normal”) and in unremarkable gastric mucosa from healthy subjects (“healthy”). A thorough investigation was carried out concerning these markers’ summary distributions, differential expression, expression correlations, as well as their relationship with patients’ clinicopathological parameters and survival outcomes.

ANXA2 emerged from our study as a protagonist in gastric adenocarcinoma oncogenesis. Its expression was significantly elevated in gastric adenocarcinoma tissues and their lymph node metastases, compared to normal and healthy controls. The gradual increase in ANXA2 expression when moving from healthy gastric tissues to normal-looking mucosa in the carcinoma vicinity, to primary adenocarcinoma and finally to lymph node metastases, hints to its role in the evolution of gastric adenocarcinomas. In addition to that, we demonstrated a significantly different ANXA2 expression between tumor grades, with higher ANXA2 levels in Grade 3 tumors compared to Grade 2, further supporting the notion that ANXA2 is linked to adverse tumor characteristics. Our results also revealed that higher levels of ANXA2 in both the primary tumors and the lymph node metastases were associated with worse overall and disease-free survival. Patients with higher ANXA2 expression in the lymph node metastases exhibited the poorest prognosis. These findings align with those of prior studies which found that ANXA2 overexpression was associated with more aggressive gastric cancer behavior and worse patient prognosis. Namely, four studies conducted in Asian populations [51–53, 56] and one in South Americans [54] showed that ANXA2 up-regulation was related to higher tumor grade, increased size, venous invasion, lymph node and distal metastasis, as well as advanced stage. The association between ANXA2 expression and gastric cancer patients’ survival was evaluated only in two Asian cohorts which reported poorer survival rates in patients with ANXA2 overexpressing gastric cancer. To the best of our knowledge, the present study is the first to highlight the dismal prognosis of ANXA2 protein overexpression in gastric adenocarcinoma in a Caucasian population.

Regarding the putative mechanism by which ANXA2 promotes gastric adenocarcinoma, some data are available. Firstly, based on patients’ tissue studies, its up-regulation was linked to reduced e-cadherin expression by Han Y. et al. [52] and to *c-erbB-2* overexpression by Emote K. et al. [56] Secondly, Tas et al. found high ANXA2 serum levels in chemotherapy-resistant patients. [60] Research in human gastric cancer cell lines demonstrated that ANXA2 inhibition reduces tumor cell migration and matrix metalloproteinases’ secretion. [53] Moreover, Leal MF et al. found that the upregulation of ANXA2 enhances gastric cancer cells invasion [54] and Zhang ZD et al. showed that its silencing reverses the tumor cells chemoresistance to cisplatin. [57] Xie R et al. confirmed the role of ANXA2 in tumor cell proliferation and survival, as well as the therapeutic potential of its silencing [58], while Mao et al. identified the EphA2–YES1–ANXA2 axis as a potential therapeutic target in gastric adenocarcinomas. [59] Finally, H. pylori seems to be able to induce ANXA2 and S100A7 overexpression, destabilizing epithelial junctions and promoting carcinogenesis. [55]

Our study reinforces the role of ANXA2 in gastric cancer tumorigenesis and progression and suggests a threshold of expression (H-score: 100) over which patients’ survival is significantly aggravated. Since protein overexpression can be easily detected in human tissues by immunohistochemistry, ANXA2 H-score may serve as a valuable prognostic biomarker. Furthermore, given that blocking its expression or disrupting its interactions with other proteins is feasible, ANXA2 emerges as a candidate therapeutic target for patients with ANXA2 overexpressing gastric adenocarcinomas. In this context, H-score could serve as a predictive marker to therapy response.

hPG, a precursor form of gastrin, has been implicated in carcinogenesis due to its involvement in various signaling pathways that promote tumor growth and survival. [24] Studies using gastric cancer cell lines and mouse models have linked hPG upregulation to aggressive tumor characteristics. More specifically, antral cells that express the progastrin receptor CCK2R have been shown to exhibit traits of stem cells. [41] hPG increases Lgr5 expression and promotes organoid formation in CCK2R+/Lgr5-cells and differentiation into Lgr5+ stem cells. [42]

Our study, which, in our knowledge, is the first to investigate hPG expression in human gastric cancer tissues by immunohistochemistry, showed some conflicting findings. One the one hand, hPG expression positively correlated with the expression of ANXA2 in the primary tumors, providing further support to their hypothesized relationship as ligand (hPG) and receptor (ANXA2), as proposed by studies investigating the possible receptors of hPG. [45,50] On the other hand, gastric cancer patients had significantly decreased hPG protein expression levels in tissue sections from the primary tumor, its lymph node metastases and the normal gastric mucosa, as compared to the healthy controls. Moreover, there were no statistically significant differences in hPG expression between the gastric tumors or their lymph node metastases and the normal gastric tissues adjacent to the tumor. Importantly, the present cohort did not reveal any significant correlation between hPG expression and the patients’ clinicopathological parameters. Our findings are partly in contrast with the sole other publication investigating hPG levels in gastric cancer patients in relation to patients’ survival. In this recent study, Amjadi O. et al. reported that increased serum levels of hPG were noted in patients with gastric cancer versus gastric-cancer free participants. However, similar to our findings, hPG levels were not significantly related to tumor prognostic features such as stage, grade, and metastatic potential. [43] Furthermore, we did not identify any statistically significant difference when comparing survival outcomes of hPG expression. Both overall and disease-free survival were unaffected by the levels of hPG in either the primary tumor or the lymph node metastases. It is of interest that investigation in other human malignancies has linked high hPG levels with worse clinical outcomes. [11] However, most of the research has been focused on serum titters [28,30,32,33,36] rather than tissue [38] hPG expression levels. Therefore, the lack of prognostic significance in our study could imply that only hPG serum concentration, as opposed to tissue protein expression, possess prognostic value. However, all data taken together, a more possible scenario is that hPG is not a reliable prognostic biomarker for gastric adenocarcinoma, indicating that the oncogenic evolution in this cancer type is more reliant on other molecular mechanisms and pathways. On the other hand, it is possible that the small sample of this specific dataset was not adequate to reveal hPG prognostic significance. It would be beneficial if future research could be conducted on a sizable set of patients to unmask any effects of hPG on gastric cancer prognosis. Another possible limitation to be considered is that there was no commercially available hPG antibody and the specificity of the one used may not have been optimal, potentially showing some cross-reactivity with other gastrins. This could also explain the lack of significant differential expression between the neoplastic and the normal-looking tissues in the current study, since gastrins are normally expressed in gastric tissue.

Tumor-associated macrophages are key components of the tumor microenvironment that can adopt different phenotypes depending on the signals they receive from the tumor milieu. The M1 phenotype is typically associated with inflammation, microbicidal and tumor suppressive activity while the M2 phenotype with tissue repair, but also with tumor promotion. [61–62] The oncogenic role of the M2 macrophage phenotype and its association with aggressive tumor features like higher grade has been demonstrated by studies on human gastric cancer tissues, including two meta-analyses. [64–68] Moreover, M2 was found to be the main TAMs phenotype in the intraperitoneal metastases of advanced human gastric cancer [69] and high levels of M2 TAMs were shown to be linked to epithelial-mesenchymal transition, both having independent negative prognostic value and being possibly linked to the TGF-β signaling pathway. [70] In addition to that, a predominance of M2 over M1 TAMs in human gastric adenoma stroma was shown to increase the likelihood of the latter transforming into gastric adenocarcinoma. [71]

In our study, the CD163/HLA-DR ratio was used to assess TAM polarization, with CD163 marking the M2 and HLA-DR the M1 macrophages. The CD163/HLA-DR ratio increased significantly as we moved from healthy to normal tissues, to primary gastric tumors and to lymph node metastases, suggesting an increasing concentration of M2 macrophages as the disease progresses. This is in line with the already mentioned studies that have shown an association between a swift in macrophages towards the M2 phenotype in lymph node metastatic disease or advanced TNM stage. It is of note that, in the present cohort, CD163/HLA-DR ratio differed significantly between tumoral and non-tumoral (normal and healthy) tissues whereas there were no statistically significant differences between normal and healthy tissues. This finding could potentially imply that microenvironment’s macrophage polarization towards an M2 phenotype is not an early event in gastric carcinogenesis, requiring accumulation of cancerous hits.

Literature supports an association between an increase in M2 TAMs and a poor patients’ survival. [64–68] In the present study, despite the observed changes in TAM polarization, the CD163/HLA-DR ratio failed to reveal any significant correlation with patient survival. In addition, the CD163/HLA-DR (T) and (LN) ratio decreased from tumor Grade 2 to 3, in contrast to previous reports that demonstrated a link between a predominance of the M2 phenotype and poor histologic differentiation. [68] Our findings are difficult to interpret and may be attributed to the small sample size of our cohort. Interestingly, our study showed a statistically significant negative correlation between ANXA2 expression and the CD163/HLA-DR ratio in tumor tissues, indicating that tumors with higher ANXA2 expression may have a lower proportion of M2 TAMs. While these results may initially seem counterintuitive given the known tumor-promoting roles of both ANXA2 and M2 macrophages, it is possible that ANXA2 overexpression triggers some compensatory mechanisms in the tumor microenvironment, including the induction of the M1 phenotype in macrophages.

While our study provides some valuable insights into the roles of hPG, ANXA2 and the TAMs in gastric adenocarcinomas, several limitations should be acknowledged. Firstly, our study was based on a relatively small sample size, which may limit the generalizability of our findings. Larger studies are needed to validate these results and to explore their potential clinical applications in gastric cancer. Additionally, as already discussed, the hPG antibody used is not a commercially available one, with a more limited documentation as to its specificity. Furthermore, while our study focused on the expression of hPG, ANXA2 and the TAMs in primary gastric tumors and their lymph node metastases, future studies should investigate their role in circulating tumor cells and distant metastases to provide a more comprehensive understanding of their role in gastric adenocarcinomas. Moreover, although we demonstrated a significant negative correlation between the expression of ANXA2 and the patients’ survival, the molecular mechanisms underlying the oncogenic effects of ANXA2 in gastric cancer remain largely unclear. Future studies should focus on elucidating the signaling pathways that are activated by ANXA2 in gastric cancer cells and how these pathways interact with other molecules or components of the tumor microenvironment. Additionally, exploring the potential therapeutic utility of targeting ANXA2, either alone or in combination with other treatments, could open new avenues for the management of gastric cancer.

## 5. Conclusion

In conclusion, the present study is the first to highlight the dismal prognosis of ANXA2 overexpression in a Caucasian population of gastric cancer patients. Our findings are also suggestive that a dichotomized ANXA2 H-score could serve as a valuable prognostic biomarker. Additionally, our work that was the first globally to investigate hPG in human gastric cancer tissues by immunohistochemistry, degrades its importance in this tumor type as no significant correlation between its expression and the patients’ clinicopathological parameters or survival outcomes was demonstrated. Finally, we verified the polarization of tumor microenvironment towards M2-like macrophages as gastric cancer progresses (Figure 7). Future research should aim to validate these findings in larger cohorts and explore the therapeutic potential of targeting ANXA2, as well as tampering with the TAM phenotype in gastric adenocarcinoma patients.

## Data Availability

All data produced in the present study are available upon reasonable request to the authors.

## Author Contributions

Conceptualization, K.C., N.C., D.S. and S.S.; Methodology, K.C., and S.S.; validation, K.C., S.M.P., R.F., N.C., N.K., D.S. and S.S.; investigation, K.C.; Formal Analysis, K.C. and S.M.P. Software, S.M.P.; writing—original draft preparation, K.C.; writing—review and editing, K.C., S.M.P., D.S., and S.S.; visualization, K.C., R.F., N.C., and S.S.; supervision, N.K., D.S. and S.S.; project administration, S.S. All authors have read and agreed to the published version of the manuscript.

## Institutional Review Board Statement

The study was conducted in accordance with the Declaration of Helsinki and approved by the Institutional Ethics Committee of Laiko General Hospital of Athens, Greece (492/18-07-2022). Individual consent was waived due to the nature of the study.

## Data Availability Statement

The data presented in this study are included in the article/supplementary material. Further inquiries can be directed to the corresponding author(s).

